# Application of Telemedicine During the Coronavirus Disease Epidemics: A Rapid Review and Meta-Analysis

**DOI:** 10.1101/2020.04.14.20065664

**Authors:** Yelei Gao, Rui Liu, Qi Zhou, Xingmei Wang, Liping Huang, Qianling Shi, Zijun Wang, Shuya Lu, Weiguo Li, Yanfang Ma, Xufei Luo, Toshio Fukuoka, Hyeong Sik Ahn, Myeong Soo Lee, Zhengxiu Luo, Enmei Liu, Yaolong Chen, Chang Shu, Daiying Tian, on behalf of COVID-19 evidence and recommendations working group

## Abstract

**Background:** As COVID-19 has become a global pandemic, early prevention and control of the epidemic is extremely important. Telemedicine, which includes medical advice given over telephone, Internet, mobile phone applications or other similar ways, may be an efficient way to reduce transmission and pressure on medical institutions.

**Methods:** We searched MEDLINE, Web of science, Embase, Cochrane, CBM, CNKI and Wanfang databases for literature on the use of telemedicine for COVID-19, SARS and MERS. from their inception to March 31st, 2020. We included studies about the content of the consultation (such as symptoms, therapy and prevention, policy, public service), screening of suspected cases, the provision of advice given to those people who may have symptoms or contact history. We conducted meta-analyses on the main outcomes of the studies.

**Results:** A total of 2041 articles were identified after removing duplicates. After reading the full texts, we finally included nine studies. People were most concerned about symptoms (64.2%), epidemic situation and public problems (14.5%), and psychological problems (10.3%) during COVID-19 epidemic. During the SARS epidemic, the proportions of people asking for consultation for symptoms, prevention and therapy, and psychological problems were 35.0%, 22.0%, and 23.0%, respectively. Two studies demonstrated that telemedicine can be used to screen the suspected patients and give advice. One study emphasized the limited possibilities to follow up people calling hotlines and difficulties in identifying all suspect cases.

**Conclusions:** Telemedicine services should focus on the issues that the public is most concerned about, such as then symptoms, prevention and treatment of the disease, and provide reasonable advice to patients with symptoms or people with epidemic history.

## Background

In December 2019, cases of pneumonia with unknown cause were broke out in China. The pathogen causing the infection was subsequently identified to be a novel coronavirus(1). On February 11th, 2020, the World Health Organization officially named the novel coronavirus pneumonia as “COVID-19”(2), and the International Committee on Taxonomy of Viruses named the novel coronavirus as “SARS-CoV-2”(3). SARS-CoV-2 can be transmitted from person to person (4-6), and the population is generally susceptible (7). Severe cases of COVID-19 are associated with acute respiratory distress syndrome, acute heart injury, shock and even death(8,9). Compared with SARS (Severe Acute Respiratory Syndrome) and MERS (Middle East Respiratory Syndrome), COVID-19 spreads faster but has lower mortality(10-12). By March 31, 2020, the virus has spread to more than 200 countries and regions in the world, with 750,890 confirmed cases and 36,405 deaths(13), resulting in a fatality among known cases as high as 4.8%. On March 11, 2020, the World Health Organization declared COVID -19 as a global pandemic.

With the rapid development of communication equipment and Internet, telemedicine has become a convenient way for the public to obtain valuable information and health consultation. Most COVID-19 patients have attended hospitals or other health facilities to be diagnosed and treated, which increases the risk of nosocomial infection(9). Remote medical treatment can reduce the unnecessary hospital visits during the outbreak and the accumulation of people in the hospital, accelerate the patients’ access to professional advice in time, and alleviate anxiousness among the members of public. Discovering, diagnosing and treating patients infected with SARS-CoV-2 as early as possible support the prevention and control of the epidemic. The purpose of this rapid review is to explore the role and potential of telemedicine during the COVID-19, SARS and MERS outbreaks.

## Methods

### Search strategy

A comprehensive search was performed by an experienced librarian in the following electronic databases from their inception to March 31st, 2020(14): the Cochrane library, MEDLINE (via PubMed), EMBASE, Web of Science, CBM (China Biology Medicine disc), CNKI (China National Knowledge Infrastructure), and Wanfang Data. We made no restrictions on language or publication status. The following search was used: (“Novel coronavirus” OR “2019-novel coronavirus” OR “Novel CoV” OR “2019-nCoV” OR “Wuhan-Cov” OR “2019-CoV” OR “Wuhan Coronavirus” OR “Wuhan seafood market pneumonia virus” OR “COVID-19” OR “SARS-CoV-2” OR “Middle East Respiratory Syndrome” OR “MERS” OR “MERS-CoV” OR “Severe Acute Respiratory Syndrome” OR “SARS” OR “SARS-CoV” OR “SARS-Related” OR “SARS-Associated”) AND (“Consultants” OR “Telemedicine” OR “Internet” OR “Counseling” OR “Consultant” OR “Consult” OR “Advisory Service” OR “Advisory Services” OR “Telehealth” OR “eHealth” OR “mHealth” OR “Mobile Health” OR “Online consultation” OR “Telephone” OR “Hotline*” OR “Online Reference Service” OR “Online Reference Desk” OR “Network Information Reference” OR “Real-time Reference Service” OR “Online inquiry” OR “Mobile Application*”OR “Mobile App*” OR “Cell Phone*” OR “Mobile phone*”). We also searched clinical trial registry platforms (the World Health Organization Clinical Trials Registry Platform (http://www.who.int/ictrp/en/), US National Institutes of Health Trials Register (https://clinicaltrials.gov/)), Google Scholar (https://scholar.google.nl/), preprint platforms (bioRxiv(https://www.biorxiv.org/),medRxiv(https://www.medrxiv.org/), SSRN (https://www.ssrn.com/index.cfm/en/)) and reference lists of the identified reviews to find unpublished and other potentially relevant studies. Finally, we contacted experts in the field to identify any relevant trials that may have been missed in our search. The details of the search strategy can be found in the **Supplementary Material 1**.

### Inclusion and exclusion criteria

We included studies that met the following criteria:1) the study population was people needing consultation related to COVID-19, SARS or MERS during the respective epidemics; and 2)the study focused on telemedicine (including the use of telephone hotline, telephone counseling, mobile application, Internet based consultations) and its use and the potential problems. There were no limitations of languages and study types. Duplicates, studies for which the full-text was unavailable, review articles, guidelines and expert consensus statements, and studies with specific data missing were excluded.

### Selection of studies

Two reviewers (R Liu and L Huang) selected the studies independently after first eliminating duplicates. The bibliographic software EndNote was used and any discrepancies were settled by discussion, consulting a third reviewer (Q Zhou) if necessary. The reviewers screened first all titles and abstracts with the pre-defined criteria, and categorized the articles into three (eligible, not eligible, and unclear) groups. In the second step, full-texts of the potentially eligible or unclear studies were reviewed to identify the final inclusion. All reasons for exclusion of ineligible studies were recorded, and the process of study selection was documented using a PRISMA flow diagram(15,16).

### Data extraction

Two reviewers (R Liu and X Wang) extracted the data independently with a standard data collection form. Disagreements were resolved by consensus, and a third reviewer(Y Gao) checked the extracted data for consistency and accuracy. Data extracted included: 1) Basic information: title, first author, publication year and study design; 2) participants: baseline characteristics and sample size; and 3) results: proportions of individuals using telemedicine for different contents of consultation (e.g. symptoms, therapy and prevention, policy, public service), details of screening of suspected cases, the provision of advice given to people who had symptoms or contact history, and the limitations of telemedicine.

### Risk of bias assessment

Two researchers (Z Wang and Q Shi) independently assessed the potential bias in each included study. The included studies were evaluated using appropriate assessment scales depending on the study type: for RCTs, the Cochrane Risk-of-Bias assessment tool, for cohort studies, the Newcastle-Ottawa Scale (NOS)(17,18), and for cross-sectional studies, the methodology evaluation tool recommended by the Agency for Healthcare Research and Quality (AHRQ)(19).

### Data synthesis

For studies on telephone hotlines, we calculated the proportions of each topic of concern among all callers, i.e. the number of individuals calling to consult on the topic under consideration divided by the total number of calls. We also collected the proportions of patients who were screened suspect case and the advice given to the caller. We did a meta-analyses of proportions, reporting the effect size (ES) with 95% confidence intervals (CI) using random-effects models(20). Two-sided *P* values < 0.05 were considered statistically significant. Heterogeneity was defined as *P*<0.10 and *I*^*2*^>50%. All analyses were performed in STATA version 14.

### Quality of the evidence assessment

Two reviewers(Z Wang and Q Shi) assessed the quality of evidence independently using the Grading of Recommendations Assessment, Development and Evaluation (GRADE) tool(21). We produced a “Summary of Findings” table using the GRADEpro software(22,23). This table shows the overall grading of body of evidence for each prespecified outcome that will be accounted for in the meta-analysis. In this approach, the quality is downgraded according to five considerations (study limitations, consistency of effect, imprecision, indirectness, and publication bias)(24) and upgraded according to three considerations (large magnitude of effect, dose-response relation and plausible confounders or biases). Finally, the quality of evidence is classified as high, moderate, low, or very low, reflecting to what extent that we are confident the effect estimates are correct.

As COVID-19 is a public health emergency of international concern and the situation is evolving rapidly, our study was not registered in order to speed up the process.(25)

## Results

### Characteristics and quality of included studies

A total of 2787 articles were identified in the database, of which 2041 articles were left after deleting the duplicates. Thirty articles were identified for further review after reviewing their titles and abstracts. Nine cross-sectional studies were finally included after reviewing the full texts(26-34). The selection process is shown in *Figure 1*. One of the nine retrieved articles was about COVID-19, and the remaining eight articles were about SARS. Eight articles assessed hotline consultation. The contents of the consultation included for instance symptoms, prevention and therapy, psychological problems, and related policies. Characteristics of the included studies were shown in *Table 1*. The quality of included studies was very poor: all studies scored less than 8 out of 11 in the evaluation by the AHRQ tool (*Table 2*).

**Table 1.** Characteristics of the included studies

| Study ID | Disease | Provider | Locations | Period | Target population | Contents of consultation | Conclusion |
| --- | --- | --- | --- | --- | --- | --- | --- |
| Li 2020(26) | COVID-19 | Internet hospital with medical consultation project team | Mianyang, Sichuan province, China | 26 Jan – 1 Feb 2020 | General public | symptoms ,psychological problems, public services, epidemic situation and public problems, other | Internet-based hospital services can screen suspected patients and carry out psychological interventions to reduce social panic and contribute to the prevention and control of the epidemic. |
| Zhang 2004 (27) | SARS | Hotline consultant | Shenyang , Liaoning province, China | 11 Apr – 21 Jun 2003 | General public | symptoms, prevention and therapy, policy, complaint and advice, epidemic situation and public problem | The consultation hotline can provide information related to the epidemic situation and reduce the psychological pressure of the social masses. |
| Zhang 2003 (28) | SARS | Hotline consultant | Beijing, China | 7 May – 10 May 2003 | General public | symptoms, prevention and therapy, psychological problem, public service, other | The fever consultation hotline can provide medical counseling services for residents, solve their fears, and save manpower and material resources. |
| Lu 2003 (29) | SARS | Hotline consultant | Shanxi province, China | 30 Apr – 9 Jun 2003 | General public | prevention and therapy, psychological problem, public service, policy, other | The counseling hotline can provide SARS prevention knowledge and psychological counseling and eliminate the panic psychology of the public. |
| Liang 2004 (30) | SARS | 24 hours Hotline consultant | Guangdong province, China | 15 Apr – 26 May 2003 | General public | symptoms, prevention and therapy, psychological problem, public service, complaint and advice, other | In catastrophic events, telephone counseling is an effective psychological intervention. |
| Li 2005 (31) | SARS | English Hotline consultant | Beijing, China | 7 Apr – 10 Jun 2003 | Foreigners | symptoms, prevention and therapy, complaint and advice, epidemic situation and public problem, other | The telephone number for foreigners consultation meets the needs of foreigners for health and disease prevention services, which is conducive to disease prevention and control. |
| Li 2005 (32) | SARS | Hotline consultant | Beijing, China | 23 Apr – 9 May 2004 | General public | symptoms, prevention and therapy, psychological problem, public service, complaint and advice, epidemic situation and public problem, other | The SARS counseling hotline provides health prevention and control and psychological counseling services for residents, which is conducive to disease prevention and control and social stability. |
| Kaydos-Daniel 2004 (33) | SARS | Fever Hotline , ( fifty-two physicians ) 8:00 a.m. -10:00 p.m. | Taiwan, Taipei | not mentioned | General public | not mentioned | Fever counseling hotline helps to find suspected SARS patients. |
| Ma 2005 (34) | SARS | Hotline consultant | Xi'an, Shanxi province, China | 18 Apr – 30 Jun 2003 | General public | symptoms , prevention and therapy, policy, complaint and advice, epidemic situation and public problem, other | The SARS hotline can provide knowledge and information about SARS. |

**Table 2.** Risk of bias in the included studies

| Study ID | Disease | 1 | 2 | 3 | 4 | 5 | 6 | 7 | 8 | 9 | 10 | 11 | Score † |
| --- | --- | --- | --- | --- | --- | --- | --- | --- | --- | --- | --- | --- | --- |
| Li 2020(26) | COVID-19 | 1 | 0 | 1 | 1 | 1 | 0 | 0 | 0 | 0 | 0 | 1 | 5 |
| Zhang 2004(27) | SARS | 1 | 0 | 1 | 1 | 0 | 0 | 0 | 0 | 0 | 0 | 1 | 4 |
| Zhang 2003(28) | SARS | 1 | 0 | 1 | 1 | 0 | 0 | 0 | 0 | 0 | 0 | 1 | 4 |
| Lu 2003(29) | SARS | 1 | 0 | 1 | 1 | 0 | 0 | 0 | 0 | 0 | 0 | 1 | 4 |
| Liang 2004(30) | SARS | 1 | 0 | 1 | 1 | 0 | 0 | 0 | 0 | 0 | 0 | 1 | 4 |
| Li 2005(31) | SARS | 1 | 0 | 1 | 1 | 0 | 0 | 0 | 0 | 0 | 0 | 1 | 4 |
| Li 2005(32) | SARS | 1 | 0 | 1 | 1 | 0 | 0 | 0 | 0 | 0 | 0 | 1 | 4 |
| Kaydos-Daniel 2004(33) | SARS | 1 | 0 | 1 | 1 | 0 | 1 | 0 | 0 | 0 | 0 | 1 | 5 |
| Ma 2005(34) | SARS | 1 | 0 | 1 | 1 | 0 | 1 | 0 | 1 | 0 | 1 | 1 | 7 |

†: According to the methodology evaluation tool recommended by the Agency for Healthcare Research and Quality. The maximum score is 1; the higher the score, the lower the risk of bias. The numbers 1 to 11 refer to the items of the tool:1) Defining the source of information (survey, record review); 2) Listing the inclusion and exclusion criteria for exposed and unexposed subjects or referring to previous publications; 3)
Indicate time period used for identifying patients; 4) Indicating whether the subjects were recruited consecutively (if not population-based); 5) Indicating if evaluators of subjective components of the study were masked from the participants; 6) Description of any assessments undertaken for quality assurance purposes (e.g., test/retest of primary outcome measurements); 7) Explaining any exclusions of patients from the analysis; 8) Description how confounding was assessed and/or controlled; 9) If applicable, explaining how missing data were handled in the analysis; 10) Summarizing patient response rates and completeness of data collection; 11) Clarification of the expected follow-up (if any), and the percentage of patients with incomplete data or follow-up.

**Figure 1.**
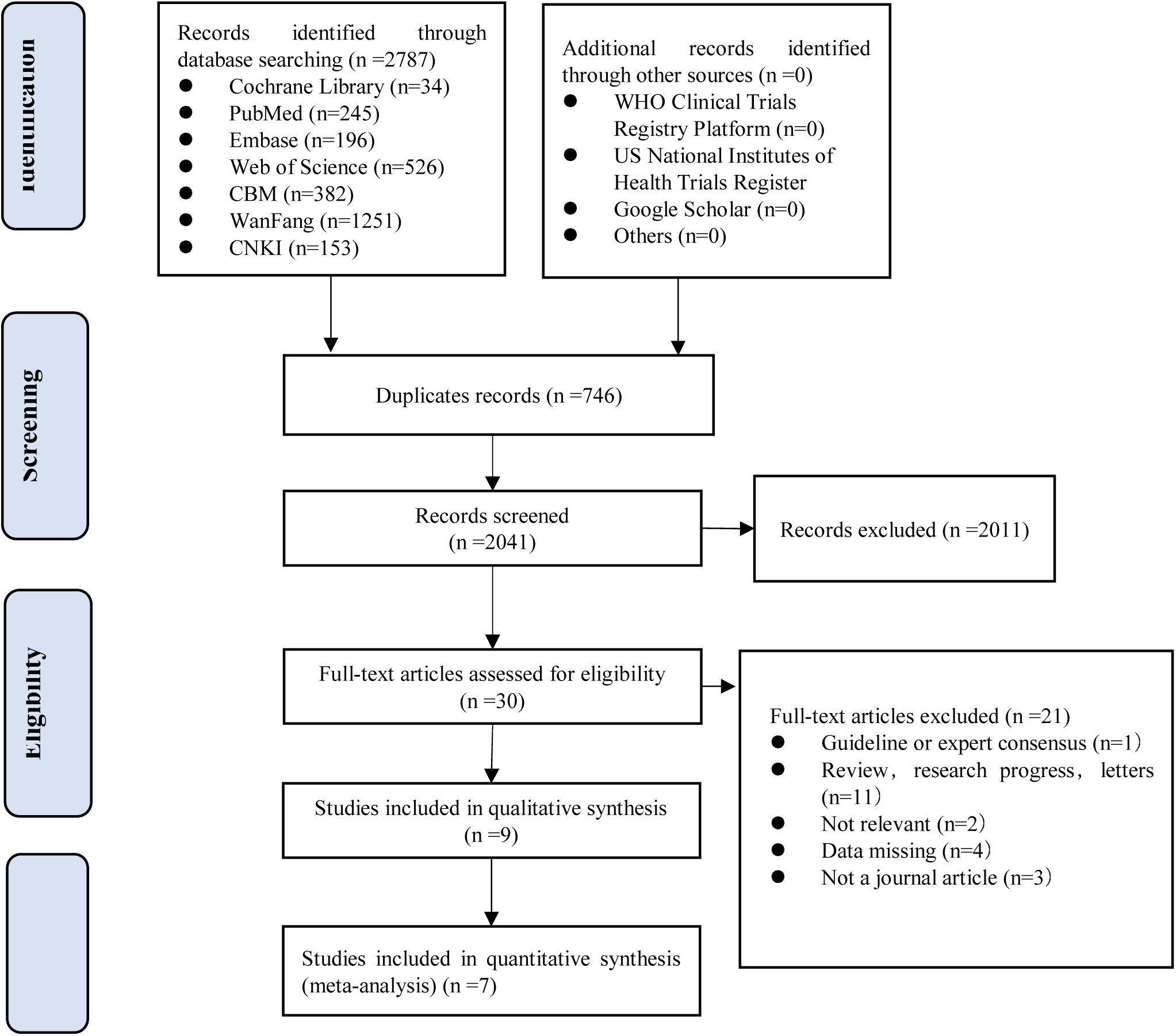
Flow diagram of the literature search.

### Contents of the consultation content

*Table 3* summarizes the quantitative findings of each study. Seven studies of SARS were conducted meta-analysis in different consultation contents (*Figure 2*).

**Table 3.** Main results of the included studies

| Study ID | Period | Number of consultations | Proportions of patients by topic of consultation (%) |  |  |  |  |  |  |  | Proportions of patients with symptoms or exposure (%) |  |  |  | Types of advice given (%) |  |  |  |
| --- | --- | --- | --- | --- | --- | --- | --- | --- | --- | --- | --- | --- | --- | --- | --- | --- | --- | --- |
|  |  |  | Symptoms | Prevention and Therapy | Psychological problem | Public service | Policy | Complaint and Advice | Epidemic situation and Public problem | Other | Fever | Exposure history | Fever and exposure history | High risk cases | No advice | Isolation at home | Go to hospital /clinics or call an ambulance | Not recorded or unclear |
| Li 2020(26) | 2020.01.26-02.01 | 4120 | 64.2 | - | 10.3 | 5.6 | - | - | 14.5 | 6.4 | 12.7 | 2.3 | 0.1 | - | 84.9 | 10.6 | 4.5 | - |
| Zhang 2004(27) | 2003.04.11-06.21 | 77313 | 59.0 | 2.0 | - | - | 13.0 | 23.0 | 4.0 | - | - | - | - | - | - | - | - | - |
| Zhang 2003(28) | 2003.05.07-05.10 | 784 | 26.7 | 31.8 | 14.9 | 21.0 | - | - | - | 5.6 | - | - | - | - | - | - | - | - |
| Lu 2003(29) | 2003.04.30-06.09 | 1370 | - | 59.9 | 60.4 | 30.7 | 23.9 | - | - | 13.9 | - | - | - | - | - | - | - | - |
| Liang 2004(30) | 2003.04.15-05.26 | 1019 | 62.7 | 16.1 | 9.4 | 4.0 | - | 0.9 | - | 6.9 | - | - | - | - | - | - | - | - |
| Li 2005(31) | 2003.04.07-06.10 | 511† | 19.4 | 23.3 | - | - | - | 11.2 | 5.6 | 40.5 | - | - | - | - | - | - | - | - |
| Li 2005(32) | 2004.04.23-05.09 | 985 | 25.5 | 13.0 | 4.0 | 10.1 | - | 3.1 | 24.2 | 18.3 | - | - | - | - | - | - | - | - |
| Kaydos-Daniel 2004(33) | 2003.06.01-06.10 | 11288 (Taiwan) | - | - | - | - | - | - | - | - | - | - | - | - | 51.0 | 21.0 | 28.0 | - |
|  |  | 1966 (Taipei) | - | - | - | - | - | - | - | - | 19.23 | - | - | 0.9 | - | 4.2 | 12.3 | 83.6 |
| Ma 2005(34) | 2003.04.18-06.30 | 14557 | 22.3 | 17.1 | - | - | - | 0.9 | 1.7 | 58.2 | - | - | - | - | - | - | - | - |

**Note :** † The total number of consultations was , but 79 were calls from the embassies to get information and thus were not included in the calculations. **Symptoms** included information of symptoms or physical signs of a disease. **Prevention and Therapy** included information on the measures to prevent infection, such as disinfection, isolation and ventilation, clinical treatments, drugs, and vaccination. **Psychological problems** included for example anxiety and fear. **Public services** included information of hospital or clinics, whether advise to travel or assembly. **Policy** included information of government policy on controlling the epidemic and insurance reimbursement. **Complaint and Advice** included complaint or tips regarding individuals suspected to be infected, or advice to the health authorities. Epidemic situation and public problem included information of the epidemic situation in the country, and public problems raised by the epidemic. Other included information not mentioned above.

**Figure 2.**
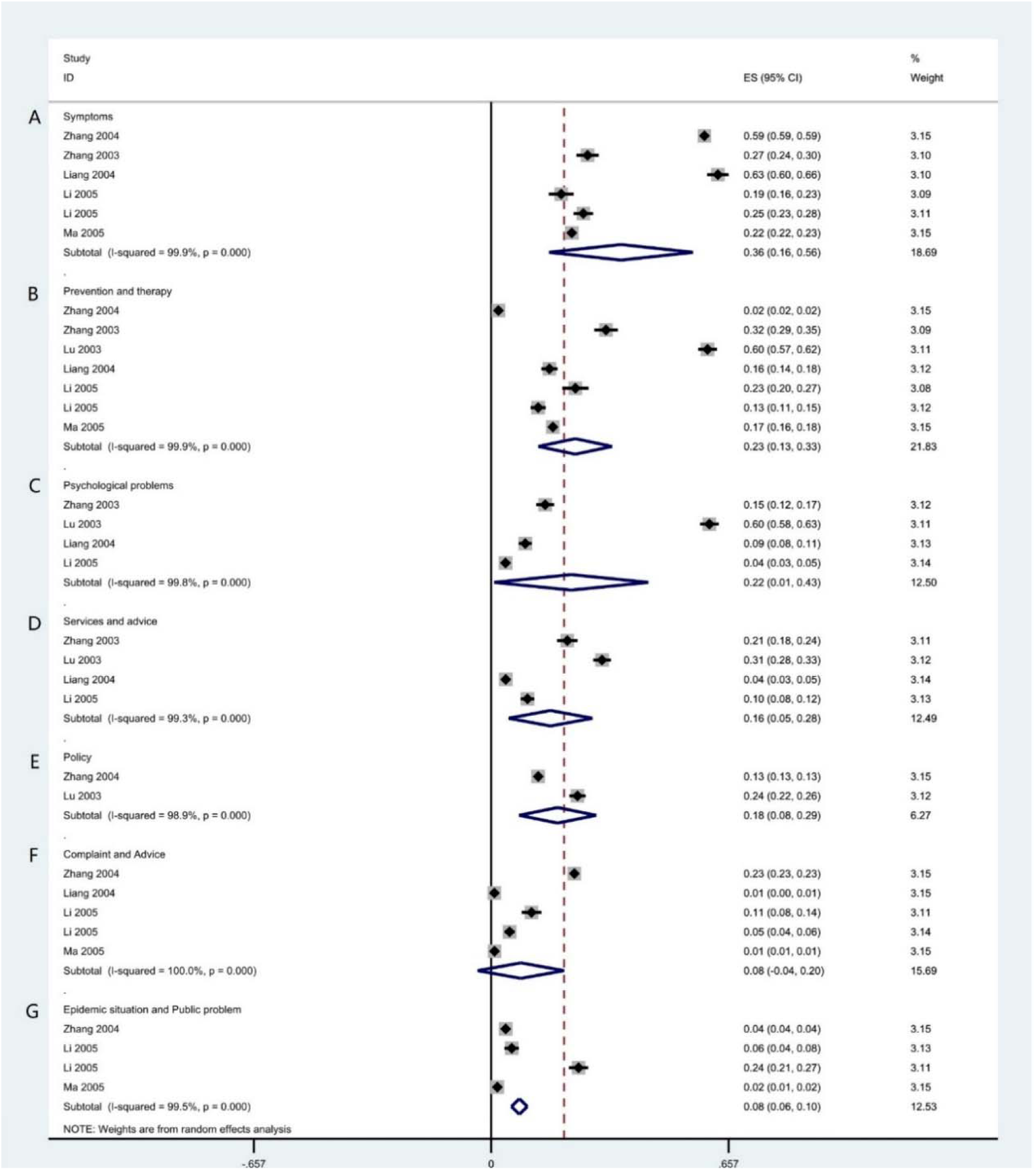
Forest plot on proportions of patients receiving consultation for different types of content of SARS.

#### Symptoms

A total of seven studies (one on COVID-19, six on SARS) reported the proportion of patients who called the hotline for counseling related to symptoms (such as fever, cough, and gastrointestinal symptoms). The proportion was 64.2% in the study conducted during the COVID-19 epidemic, and 36.0%, 95% CI: 0.16 to 0.56 (*I*^*2*^ = 99.9%) in the studies during the SARS epidemic (*Figure 2A*).

#### Prevention and therapy

Seven studies reported the proportions of patients counseled for prevention and treatment (including disinfection, isolation, ventilation, drug treatment, and vaccination). All seven studies were about SARS. The pooled proportion was 23.0%, 95% CI: 0.13 to 0.33 (*I*^*2*^ = 99.9%) (*Figure 2B*).

#### Psychological problems

Five studies (one on COVID-19, four on SARS) reported the proportion of patients receiving psychological counseling (counseling content includes anxiety, worry and fear of infection related to COVID-19 or SARS). The proportion of psychological consultation is 10.3% in the COVID-19 study and 22.0% in the SARS studies, 95% CI: 0.01 to 0.43 (*I*^*2*^ = 99.8%) (*Figure 2C*).

#### Services and advice

Five studies (including one on COVID-19) reported the proportion of patients receiving consultation about services and advice (such as which hospital to visit if suspecting coronavirus infection, whether it is advisable to travel or organize meetings or events. The proportion was 5.6% during in the COVID-19 study and was 16.0%, 95% CI: 0.05 to 0.28 (*I*^*2*^ = 99.3%) in the SARS studies (*Figure 2D*).

#### Policy

Two studies, both on SARS, reported the proportion of patients receiving policy consultations (meaning e.g. local SARS prevention and control policies, SARS medical insurance reimbursement policies, or hospitalization expenses). The respective pooled proportion was 18.0%, 95% CI: 0.08 to 0.29 (*I*^*2*^ = 98.9%) (*Figure 2E*).

#### Complaint and Advice

A total of five studies focused on the proportion of reported complaints and advice (consultation content includes for example reporting suspected patients, or people with close contact with confirm case, and complaints of policy implementation). All the five studies were related to SARS. The results of meta-analysis of the random effect model show that the proportion of reporting and complaint consultation is 8.0%, 95% CI: 0.00 to 0.20 (*I*^*2*^ = 100.0%) (*Figure 2F*).

### Epidemic situation and Public problem

Five studies (including one on COVID-19) assessed the epidemic situation and the proportion of consultation on public issues (including disease knowledge, epidemic situation and public issues of COVID-19/SARS). The respective proportion was 14.5% in the COVID-19 study, and 8.0% in the studies conducted during the SARS epidemic (95% CI: 0.06 to 0.10; *I*^*2*^ = 99.5%) (*Figure 2G*).

### Using telemedicine to initially screen patients for suspected coronavirus infection

Two studies assessed the screening of COVID-19 or SARS. The study on COVID-19 showed that among 4,120 people who needed consultation, 524 had fever, 93 had a history of exposure though contacts; five patients had both fever and contact exposure. The other study on SARS from Taipei showed that of the 1966 patients, 378 had fever, and 18 were considered to be at high risk of having SARS.

### Advice provision by telemedicine

Two studies assessed the provision of advice for patients with COVID-19 or SARS given by medical experts. The study on COVID-19 showed that 437 (10.6%) of the 4120 patients were advised to stay at home for medical observation and 185 (4.5%) were advised to go to a hospital or clinic, or call an ambulance. The other study on SARS showed that of the 11288 patients, 21.0% were advised to isolate at home for observation, and 28.0% were advised to go to the clinic, hospital or call an ambulance.

### Limitations of telemedicine

One studies of SARS pointed out that because people were not followed up for outcomes and hotline data were not collected systematically, it impossible to determine how many of the patients who were suspected to be at risk of having SARS based on the telemedicine consultation were subsequently tested.

### Quality of evidence

The results of GRADE on main outcomes showed that the quality of evidence on consultation contents were low or very low. The details can be found in the Supplementary Material 2.

## Discussion

Our study showed that among the people who need consultation, COVID-19 patients were most concerned about symptoms, the epidemic situation and public problems related to the disease, whereas SARS patients were most concerned about symptoms, prevention and treatment, and psychological problems. Internet based health services and telephone hotlines were also shown to be able to initially identify some suspected patients and provide medical advice. Internet based health services and telephone hotlines can help to identify people with fever, exposure history or high-risk and make suggestions depending on their condition. The consultants providing telemedicine services were usually medical experts(26,33).

According to the current knowledge, SARS-CoV-2 is mainly transmitted by droplets and direct contact, and family clusters and nosocomial infection are also common(9,35,36).It is important to reduce transmission as much as possible. During infectious diseases epidemics, the public can efficiently get advice and assessment for the disease through telephone hotlines, other telemedicine services and online hospital services. This way the risk of exposing of uninfected people can also be mitigated.

We found that people were not only concerned about the disease itself, but they also needed other kinds of help during the epidemics. People who are isolated because of an infectious disease may develop negative psychological reactions, such post-traumatic stress symptoms, confusion, anger, and fear of infection(37). Telephone hotlines can be used as a tool for psychological interventions after the outbreak of the epidemic(30), so that people can obtain the necessary knowledge and a platform to relieve their anxiety and fear.

During the outbreak of COVID-19, hospitals have implemented outpatient pharmaceutical care using an Internet based medical care model. This model helps to provide medication for patients with chronic diseases without them needing to leave home, and ensure the sustainability of medication for patients with chronic diseases and reduce the risk of potential cross-infection. Internet-based medical care also has been shown to save nearly 3.5 hours of time for prescribing medicine during follow-up, 1.9 hours of time spent in hospital, and about 55.6 CNY of travel and meal expenses for per patient(38) on 1^st^ to 7^th^ February, in West China Hospital of Sichuan University during the epidemic.

Telephone and Internet services are being widely used for influenza counselling and surveillance(39-41). According to a prospective study on influenza, a self-triage Internet based decision-making tool could help parents and adult caregivers to determine when children with influenza-like illnesses need to go to the emergency department. Fourteen of the 15 patients who eventually needed to go to the emergency department were classified as high risk by this tool, resulting in a sensitivity of 93.3% (95% CI 68.1% to 99.8%). An Internet based self-triage tool can thus be feasible(42). Self-triage may be an effective way to encourage appropriate practice and reduce the pressure on health system services. Another study also showed that combining telephone and Internet services can be used by primary care facilities to promote patient self-management during flu season(43),and provided patients with medical advice and oseltamivir prescriptions(39).

Although telemedicine can be applied to the early prevention and control of infectious diseases, there are still some deficiencies. For example, one study from Taiwan on SARS pointed out that the lack of systematic data collection by the telemedicine hotlines meant that it is not possible to know whether all callers who were identified as being at high risk of having SARS eventually followed their recommendations and went to the medical facility(33). Some hotlines are open 24 hours a day, meaning that it may be difficult to find operators with sufficient professional knowledge. If the operators do not have enough professional knowledge, they may provide wrong information misleading the public, or provide inappropriate medical advice, leading to a treatment delay or missed diagnoses.

From these studies, we can conclude that telemedicine on one hand provides the public with access to medical resources or information, improves the awareness of diseases, and relieves psychological stress. On the other hand, it helps to protect the privacy of the patients, prevents people from going unnecessarily to hospitals and clinics, prevents nosocomial infection, and reduces the pressure on medical institutions. Telemedicine also helps to get feedback that can be used to make decisions.

### Strengths and limitations

This study is to our knowledge the first systematic and comprehensive exposition of telemedicine consultation during the COVID-19, SARS and MERS epidemics. Our study had however several limitations. We found no original studies comparing telemedicine with traditional medical services in the prevention and control of COVID-19. We also did not find any studies about telemedicine use during the MERS outbreak. In the future, more research is needed to evaluate the role of telemedicine in acute infectious diseases. It is also necessary to establish a reliable telemedicine service system as soon as possible after an outbreak of an infectious disease, to help distinguish the patients with the emerging infection from other patients.

## Conclusion

Telemedicine offers the public an efficient and safe way to consult healthcare professionals about the symptoms of infectious diseases, prevention and treatment measures, public health services, psychological, and other issues that the public are mostly concerned about. This way, the public can access the medical information conveniently and quickly, and reduce the risk of exposure to the infection within hospitals or clinics. Telemedicine can help in the screening of suspected infectious disease patients, which can prevent and control early infection, reduce the spread of SARS-CoV-2, and reduce the burden of medical staff in medical institutions.

## Data Availability

This study is a review.All data were from the published original studies and real.

## Author contributions

(I) Conception and design: Y Chen and E Liu; (II) Administrative support: Y Chen; (III) Provision of study materials or patients: Y Gao, R Liu and X Wang; (IV) Collection and assembly of data: R Liu and X Wang; (V) Data analysis and interpretation: Q Zhou, Y Gao, Q Shi and Z Wang; (VI) Manuscript writing: All authors; (VII) Final approval of manuscript: All authors.

## Acknowledgments

We thank Janne Estill, Institute of Global Health of University of Geneva for providing guidance and comments for our review. We thank all the authors for their wonderful collaboration.

## Footnote

### Ethical Statement

The authors are accountable for all aspects of the work in ensuring that questions related to the accuracy or integrity of any part of the work are appropriately investigated and resolved.

## Supplementary Material 1-Search strategy

### PubMed

#1 “COVID-19”[Supplementary Concept]

#2 “Severe Acute Respiratory Syndrome Coronavirus 2”[Supplementary Concept]

#3 “Middle East Respiratory Syndrome Coronavirus”[Mesh]

#4 “Severe Acute Respiratory Syndrome”[Mesh]

#5 “SARS Virus”[Mesh]

#6 “COVID-19”[Title/Abstract]

#7 “SARS-COV-2”[Title/Abstract]

#8 “Novel coronavirus” [Title/Abstract]

#9 “2019-novel coronavirus” [Title/Abstract]

#10 “coronavirus disease-19” [Title/Abstract]

#11 “coronavirus disease 2019” [Title/Abstract]

#12 “COVID19” [Title/Abstract]

#13 “Novel CoV” [Title/Abstract]

#14 “2019-nCoV” [Title/Abstract]

#15 “2019-CoV” [Title/Abstract]

#16 “Wuhan-Cov” [Title/Abstract]

#17 “Wuhan Coronavirus” [Title/Abstract]

#18 “Wuhan seafood market pneumonia virus” [Title/Abstract]

#19 “Middle East Respiratory Syndrome” [Title/Abstract]

#20 “MERS” [Title/Abstract]

#21 “MERS-CoV” [Title/Abstract]

#22 “Severe Acute Respiratory Syndrome” [Title/Abstract]

#23 “SARS” [Title/Abstract]

#24 “SARS-CoV” [Title/Abstract]

#25 “SARS-Related” [Title/Abstract]

#26 “SARS-Associated” [Title/Abstract]

#27 #1-#26/ OR

#28 “Consultant” [MeSH Terms]

#29 “Telemedicine” [MeSH Terms]

#30 “Internet” [MeSH Terms]

#31 “Mobile Applications” [MeSH Terms]

#32 “Counseling” [MeSH Terms]

#33 “Consultant*” [Title/Abstract]

#34 “Consult” [Title/Abstract]

#35 “Advisory Service*” [Title/Abstract]

#36 “Telehealth” [Title/Abstract]

#37 “eHealth” [Title/Abstract]

#38 “mHealth” [Title/Abstract]

#39 “Mobile Health” [Title/Abstract]

#40 “Online consultation” [Title/Abstract]

#41 “telephone” [Title/Abstract]

#42 “hotline*” [Title/Abstract]

#43 “counseling” [Title/Abstract]

#44 “Online Reference Service” [Title/Abstract]

#45 “Online Reference Desk” [Title/Abstract]

#46 “Network Information Reference” [Title/Abstract]

#47 “Real-time Reference Service” [Title/Abstract]

#48 “Online inquiry” [Title/Abstract]

#49 “Mobile Application*” [Title/Abstract]

#50 “Mobile App*” [Title/Abstract]

#51 “Cell Phone*” [Title/Abstract]

#52 “Mobile phone*” [Title/Abstract]

#53 “Internet*” [Title/Abstract]

#54 #28-# 53/OR

#55 #27 AND #54

### Embase

#1 ‘middle east respiratory syndrome coronavirus’/exp

#2 ‘severe acute respiratory syndrome’/exp

#3 ‘sars coronavirus’/exp

#4 ‘COVID-19’:ab,ti

#5 ‘SARS-COV-2’:ab,ti

#6 ‘novel coronavirus’:ab,ti

#7 ‘2019-novel coronavirus’:ab,ti

#8 ‘coronavirus disease-19’:ab,ti

#9 ‘coronavirus disease 2019’:ab,ti

#10 ‘COVID19’:ab,ti

#11 ‘novel cov’:ab,ti

#12 ‘2019-ncov’:ab,ti

#13 ‘2019-cov’:ab,ti

#14 ‘wuhan-cov’:ab,ti

#15 ‘wuhan coronavirus’:ab,ti

#16 ‘wuhan seafood market pneumonia virus’:ab,ti

#17 ‘middle east respiratory syndrome’:ab,ti

#18 ‘middle east respiratory syndrome coronavirus’:ab,ti

#19 ‘mers’:ab,ti

#20 ‘mers-cov’:ab,ti

#21 ‘severe acute respiratory syndrome’:ab,ti

#22 ‘sars’:ab,ti

#23 ‘sars-cov’:ab,ti

#24 ‘sars-related’:ab,ti

#25 ‘sars-associated’:ab,ti

#26 #1-#25/ OR

#27 ‘telemedicine’:ab,ti

#28 ‘mobile applications’:ab,ti

#29 ‘consultant*’:ab,ti

#30 ‘consult’:ab,ti

#31 ‘advisory service’:ab,ti

#32 ‘advisory services’:ab,ti

#33 ‘telehealth’:ab,ti

#34 ‘ehealth’:ab,ti

#35 ‘mhealth’:ab,ti

#36 ‘mobile health’:ab,ti

#37 ‘online consultation’:ab,ti

#38 ‘telephone’:ab,ti

#39 ‘hotline*’:ab,ti

#40 ‘counseling’:ab,ti

#41 ‘online reference service’:ab,ti

#42 ‘online reference desk’:ab,ti

#43 ‘network information reference’:ab,ti

#44 ‘real-time reference service’:ab,ti

#45 ‘online inquiry’:ab,ti

#46 ‘mobile application*’:ab,ti

#47 ‘mobile app*’:ab,ti

#48 ‘cell phone*’:ab,ti

#49 ‘mobile phone*’:ab,ti

#50 ‘internet:ab,ti

#51 #27-#50/ OR

#52 #26 AND #51

### Web of science

#1 TOPIC: “COVID-19”

#2 TOPIC: “SARS-COV-2”

#3 TOPIC: “Novel coronavirus”

#4 TOPIC: “2019-novel coronavirus”

#5 TOPIC: “coronavirus disease-19”

#6 TOPIC: “coronavirus disease 2019”

#7 TOPIC: “COVID 19”

#8 TOPIC: “Novel CoV”

#9 TOPIC: “2019-nCoV”

#10 TOPIC: “2019-CoV”

#11 TOPIC: “Wuhan-Cov”

#12 TOPIC: “Wuhan Coronavirus”

#13 TOPIC: “Wuhan seafood market pneumonia virus”

#14 TOPIC: “Middle East Respiratory Syndrome”

#15 TOPIC: “MERS”

#16 TOPIC: “MERS-CoV”

#17 TOPIC: “Severe Acute Respiratory Syndrome”

#18 TOPIC: “SARS”

#19 TOPIC: “SARS-CoV”

#20 TOPIC: “SARS-Related”

#21 TOPIC: “SARS-Associated”

#22 #1-#21/OR

#23 TOPIC: “Online consultation”

#24 TOPIC: “mobile Health”

#25 TOPIC: “mHealth”

#26 TOPIC: “eHealth”

#27 TOPIC: “Telehealth”

#28 TOPIC: “Advisory Service*”

#29 TOPIC: “Consult”

#30 TOPIC: “Counseling”

#31 TOPIC: “Mobile Applications”

#32 TOPIC: “Internet”

#33 TOPIC: “Telemedicine”

#34 TOPIC: “Consultant*”

#35 TOPIC: “Hotline*”

#36 TOPIC: “Telephone”

#37 TOPIC: “Counseling”

#38 TOPIC: “Online Reference Service”

#39 TOPIC: “Online Reference Desk”

#40 TOPIC: “Network Information Reference”

#41 TOPIC: “Real-time Reference Service”

#42 TOPIC: “Online inquiry”

#43 #23-#42/OR

#44 #22 AND #43

### Cochrane library

#1 MeSH descriptor: [Middle East Respiratory Syndrome Coronavirus] explode all trees

#2 MeSH descriptor: [Severe Acute Respiratory Syndrome] explode all trees

#3 MeSH descriptor: [SARS Virus] explode all trees

#4 “COVID-19”:ti,ab,kw

#5 “SARS-COV-2”:ti,ab,kw

#6 “Novel coronavirus”:ti,ab,kw

#7 “2019-novel coronavirus” :ti,ab,kw

#8 “Novel CoV” :ti,ab,kw

#9 “2019-nCoV” :ti,ab,kw

#10 “2019-CoV” :ti,ab,kw

#11 “coronavirus disease-19” :ti,ab,kw

#12 “coronavirus disease 2019” :ti,ab,kw

#13 “COVID19” :ti,ab,kw

#14 “Wuhan-Cov” :ti,ab,kw

#15 “Wuhan Coronavirus” :ti,ab,kw

#16 “Wuhan seafood market pneumonia virus” :ti,ab,kw

#17 “Middle East Respiratory Syndrome” :ti,ab,kw

#18 “MERS”:ti,ab,kw

#19 “MERS-CoV”:ti,ab,kw

#20 “Severe Acute Respiratory Syndrome”:ti,ab,kw

#21 “SARS” :ti,ab,kw

#22 “SARS-CoV” :ti,ab,kw

#23 “SARS-Related”:ti,ab,kw

#24 “SARS-Associated”:ti,ab,kw

#25 #1-#24/ OR

#26 “online reference desk”:ti,ab,kw

#27 “online reference service”:ti,ab,kw

#28 “counseling”:ti,ab,kw

#29 “hotline*”:ti,ab,kw

#30 “telephone”:ti,ab,kw

#31 “online consultation”:ti,ab,kw

#32 “mobile health”:ti,ab,kw

#33 “mhealth”:ti,ab,kw

#34 “ehealth”:ti,ab,kw

#35 “telehealth”:ti,ab,kw

#36 “advisory service*”:ti,ab,kw

#37 “consult”:ti,ab,kw

#38 “consultant*”:ti,ab,kw

#39 “mobile applications”:ti,ab,kw

#40 “telemediciner”:ti,ab,kw

#41 “network information reference”:ti,ab,kw

#42 “real-time reference service”:ti,ab,kw

#43 “mobile application*”:ti,ab,kw

#44 “internet”:ti,ab,kw

#45 “online inquiry”:ti,ab,kw

#46 “mobile phone*”:ti,ab,kw

#47 “cell phone*”:ti,ab,kw

#48 “mobile app*”:ti,ab,kw

#49 #26-#48/OR

#50 #25 AND #49

### CNKI

#1 “新型冠状病毒”[主题]

#2 “COVID-19”[主题]

#3 “COVID 19”[主题]

#4 “2019-nCoV”[主题]

#5 “2019-CoV”[主题]

#6 “SARS-CoV-2”[主题]

#7 “武汉冠状病毒”[主题]

#8 “中东呼吸综合征”[主题]

#9 “MERS”[主题]

#10 “MERS-CoV”[主题]

#11 “严重急性呼吸综合征”[主题]

#12 “SARS”[主题]

#13 #1-#12/ OR

#14 “热线”[主题]

#15 “咨询”[主题]

#16 “移动医疗”[主题]

#17 “远程医疗”[主题]

#18 “电话”[主题]

#19 “网络”[主题]

#20 ‘“因特网”[主题]

#21 “互联网”[主题]

#22 “手机应用程序”[主题]

#23 #14-#22/ OR

#24 #13 AND #23

### WanFang

#1 “新型冠状病毒”[主题]

#2 “COVID-19”[主题]

#3 “COVID 19”[主题]

#4 “2019-nCoV”[主题]

#5 “2019-CoV”[主题]

#6 “SARS-CoV-2”[主题]

#7 “武汉冠状病毒”[主题]

#8 “中东呼吸综合征”[主题]

#9 “MERS”[主题]

#10 “MERS-CoV”[主题]

#11 “严重急性呼吸综合征”[主题]

#12 “SARS”[主题]

#13 #1-#12/ OR

#14 “移动医疗”[主题]

#15 “远程医疗”[主题]

#16 “电话”[主题]

#17 “因特网”)[主题]

#18 “因特网”)[主题]

#19 “互联网”[主题]

#20 “咨询”[主题]

#21 “手机应用程序”[主题]

#22 “热线”[主题]

#23 #14-#22/OR

#24 #18 AND #23

### CBM

#1 “新型冠状病毒”[常用字段:智能]

#2 “COVID-19”[常用字段:智能]

#3 “COVID 19”[常用字段:智能]

#4 “2019-nCoV”[常用字段:智能]

#5 “2019-CoV”[常用字段:智能]

#6 “SARS-CoV-2”[常用字段:智能]

#7 “武汉冠状病毒”[常用字段:智能]

#8 “中东呼吸综合征冠状病毒”[不加权:扩展]

#9 “中东呼吸综合征”[常用字段:智能]

#10 “MERS”[常用字段:智能]

#11 “MERS-CoV”[常用字段:智能]

#12 “严重急性呼吸综合征”[不加权:扩展]

#13 “SARS 病毒”[不加权:扩展]

#14 “严重急性呼吸综合征”[常用字段:智能]

#15 “SARS”[常用字段:智能]

#16 #1-#15/OR

#17 “移动医疗”[常用字段:智能]

#18 “远程医疗”[常用字段:智能]

#19 “网络”[常用字段:智能]

#20 “电话”[常用字段:智能]

#21 “因特网”[常用字段:智能]

#22 “互联网”[常用字段:智能]

#23 “咨询&#x201D;[常用字段:智能]

#24 “手机应用程序”[常用字段:智能]

#25 “热线”[常用字段:智能]

#26 #17-#25/OR

#27 #16 AND #26

## Supplementary Material 2-Summary of Findings

| № of studies | Sample Size | Certainty assessment |  |  |  |  | Effect Value (95% CI) | Certainty |
| --- | --- | --- | --- | --- | --- | --- | --- | --- |
|  |  | Risk of bias | Inconsistency | Indirectness | Imprecision | Other considerations |  |  |
| **Symptoms** |  |  |  |  |  |  |  |  |
| 6 CSs | 95169 | serious<sup>1</sup> | serious<sup>2</sup> | not serious | serious<sup>3</sup> | none | 36% (16%, 56%) | ⊕○○○<br>VERY LOW |
| **Prevention and therapy** |  |  |  |  |  |  |  |  |
| 7 CSs | 96539 | serious<sup>1</sup> | serious<sup>2</sup> | not serious | not serious | none | 23% (13%, 33%) | ⊕⊕○○<br>LOW |
| **Psychological problems** |  |  |  |  |  |  |  |  |
| 4 CSs | 4158 | serious<sup>1</sup> | serious<sup>2</sup> | not serious | serious<sup>3</sup> | none | 22% (1%, 43%) | ⊕○○○<br>VERY LOW |
| **Services and advice** |  |  |  |  |  |  |  |  |
| 4 CSs | 4158 | serious<sup>1</sup> | serious<sup>2</sup> | not serious | not serious | none | 16% (5%, 28%) | ⊕⊕○○<br>LOW |
| **Policy** |  |  |  |  |  |  |  |  |
| 2 CSs | 78683 | serious<sup>1</sup> | serious<sup>2</sup> | not serious | not serious | none | 18% (8%, 29%) | ⊕⊕○○<br>LOW |
| **Complaint and Advice** |  |  |  |  |  |  |  |  |
| 5 CSs | 94385 | serious<sup>1</sup> | serious<sup>2</sup> | not serious | not serious | none | 8% (0%, 20%) | ⊕⊕○○<br>LOW |
| **Epidemic situation and Public problem** |  |  |  |  |  |  |  |  |
| 4 CSs | 93366 | serious<sup>1</sup> | serious<sup>2</sup> | not serious | not serious | none | 8% (6%, 10%) | ⊕⊕○○<br>LOW |

CI: Confidence Interval; CS: Cross-sectional study;

